# Polymorphism *Q223R* of the leptin receptor (*LEPR*) - a possible relationship with adaptation to non-tropical climate in Yakuts

**DOI:** 10.1101/2023.10.09.23296771

**Authors:** N.I. Pavlova, A.A. Bochurov, V.A. Alekseev, A.V. Krylov, L.A. Sydykova, Kh.A. Kurtanov

## Abstract

Obesity is an energy imbalance that occurs due to a lack of energy intake and consumption. We studied the variability of the *Q223R* polymorphism of the *LEPR* gene in the Yakut population and the relationship with body mass index (BMI) and abdominal obesity in a sample of Yakuts (n=336), consisting of individuals with obesity (n=185) and normal weight (n=151). For genotyping, we used the classical methods of PCR-RFLP analysis. A comparative analysis of the obtained data on the frequencies of alleles and genotypes with data on other populations of the world was also carried out. The G variant allele frequency was 79.5% in normal weight patients and 82.7% in obese patients. Genotype analysis showed a high frequency of genotypes GG - 64.2% and GA - 30.5% in the group with normal BMI and GG - 69.7% and GA - 25.9% in the group with high BMI. There was no significant difference in the frequency of alleles and genotypes of the *Q223R* polymorphism between the groups. It was established that the frequency of the G allele of the Yakuts (79.5%) with the populations of East Asia (86.9%). When analyzing the average anthropometric values, depending on the genotype, a statistically significant difference in waist circumference was found in persons with abdominal obesity (p = 0.03), so it was greater in carriers of the heterozygous AG genotype than in carriers of the GG genotype.

In conclusion, our study demonstrates that SNP *Q223R* (*LEPR*) is possible and has some effect on anthropometric parameters in the Yakut population, but differs from studies conducted on samples of European ethnicity. It can be assumed that the accumulation of the G allele of the *Q223R* polymorphism (*LEPR*) in the Yakut population, as well as in the populations of East Asia, is probably the result of metabolic adaptation to living conditions in a non-tropical climate.

## Introduction

Over the past few decades, the prevalence and incidence of obesity have increased rapidly worldwide and reached epidemic proportions. Obesity is associated with many adverse consequences, such as type 2 diabetes, hypercholesterolemia, hypertension or heart disease, and is directly associated with increased mortality and reduced life expectancy [1]. With the completion of the Human Genome Project and the first large genome-wide association studies, an increasing number of risk alleles associated with obesity have been identified, some in genes not previously known to be associated with obesity. However, to date, genome-wide association studies have identified only a small percentage of genetic variation significantly associated with obesity or body mass index for most ethnic groups.

The mechanisms and genetic basis of the influence of diet and habitat remain unclear. One widely studied candidate gene for obesity, the leptin receptor (*LEPR*) gene, is located in the biological pathway to obesity (leptin-insulin pathway). Leptin is produced in adipose tissue in proportion to its mass and in other organs. It is known to have pleiotropic effects, including the regulation of several neuropeptides involved in appetite control and thermogenesis. Numerous studies have tested two nonsynonymous single nucleotide polymorphisms of the *LEPR* gene (*Q223R* and *K109R*) for association with obesity and type 2 diabetes, with inconclusive results [2, 3]. Recently, many studies have been published on the association between LEPR variants and obesity, including studies of the interaction of these variants with sex or other factors. Of these two polymorphisms, the *Q223R* polymorphism results from a nonconservative A to G substitution at codon 223, resulting in a change of the amino *acRid* glutamine to arginine. This functional variant reduces leptin binding and thus impairs leptin signaling. Data from studies of the *Q223R* polymorphism are very contradictory, taking into account the results obtained in patients from different ethnic groups, which makes further studies of polymorphic loci of the *LEPR* gene relevant.

The purpose of this study was to conduct a comparative analysis of the frequencies of alleles and genotypes of the *Q223R* polymorphism of the *LEPR* gene in the Yakut population in samples with normal weight and obesity, and also to compare the data obtained with other populations in the world.

## Materials and methods

### Participants

The experimental part of the study on genotyping the *Q223R* polymorphism of the *LEPR* gene was carried out in the laboratory of hereditary pathology of the department of molecular genetics of the Yakut Scientific Center for Complex Medical Problems (YSC CMP). The research material was DNA samples from the collection of biomaterial (DNA) of populations of the Republic of Sakha (Yakutia) YSC CMP, using the Unique scientific equipment (USE) “The Genome of Yakutia” (No. USU_507512). The study included participants who filled out a questionnaire approved by the Local Committee on Biomedical Ethics at the YSC CMP and voluntarily signed an informed consent to conduct a genetic study.

A total of 336 DNA samples from volunteers without chronic diseases (117 women and 219 men) of Yakut nationality, whose average age was 47.4 ± 0.06 years, were studied. Two groups of subjects were formed: a group with normal BMI (n=151), a group with obesity (n=185). In accordance with WHO recommendations, obesity was diagnosed with a BMI > 30 kg/m2; indicators from 18.0 to 24.9 kg/m2 were considered normal. Abdominal obesity was diagnosed when the waist circumference exceeded 88 cm in women and 102 cm in men.

### Genetic Analyses

To carry out molecular genetic analysis, genomic DNA samples were isolated from whole blood using a commercial DNA isolation kit Newteryx (Yakutsk, Russia). Single nucleotide polymorphism was determined by polymerase chain reaction (PCR) followed by restriction fragment length polymorphism (RFLP) analysis. Amplification of the gene region containing the polymorphic variant was carried out with standard pairs of primers Forward: 5′-ACCCTTTAAGCTGGGTGTCCCAAATAG-3 and Reverse: 5′ - AATGTCAGTTCAGCCCATAAATATGG -3′. PCR temperature conditions were as follows: 94°C for 4 min, followed by 35 cycles at 94°C for 1 min, 62°C for 1 min, and 72°C for 1 min, and a final extension at 72°C for 5 min. The RFLP mixture with a volume of 20 μl consisted of: amplifier - 7 μl, deionized water - 10.9 μl, restriction buffer - 2 μl and restriction endunuclease MspI (2 u.a.). Interpretation of the genotyping results was performed based on different patterns of gene region bands (Fig 1).

**Fig 1.**
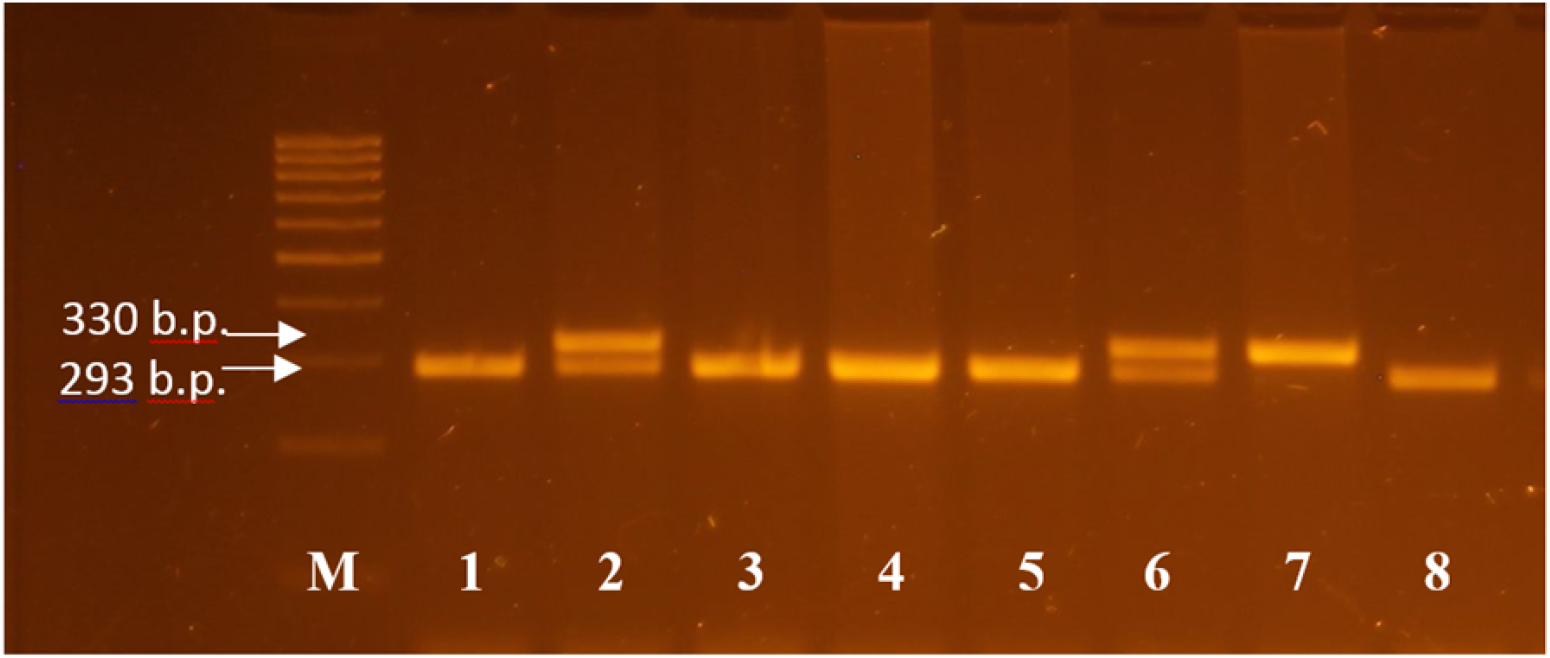
Electropherogram of the *LEPR* gene region in a 4% agarose gel after RFLP. 1 - Step 100 marker; 7 – genotype AA (330 bp); 2 and 6 – genotype AG (330, 293 bp); 1, 3,4,5,8 – genotype GG (293 bp).

### Statistical analyses

Statistical analysis of the research results was carried out using the program: “Office Microsoft Excel 2010”, “Statistica 8.0”. When analyzing the association between the frequency of an unfavorable allele and obesity, a four-field contingency table and the χ-square test with Yates’ correction were used. To assess the significance of the odds ratio, the boundaries of the 95% confidence interval (CI 95%) were calculated. To calculate the odds ratio, the following formula was used: OR = “A*D” / “B*C” (where OR is the odds ratio; A, B, C, D is the number of observations in the cells of the contingency table). To assess the normality of the sample distribution, the Kolmogorov–Smirnov test was used, which indicated a distribution different from normal. Pairwise comparison of mean BMI values depending on genotype was carried out using the Mann–Whitney test. Results were considered significant at p<0.05.

## Results

The variability of the *Q223R* polymorphism of the *LEPR* gene in the Yakut population and the effect on obesity (abdominal obesity) in Yakuts with high and normal BMI were studied. The study sample consisted of 336 patients of Yakut nationality. Overweight or obesity (BMI>30 kg/m2) was observed in 185 (55%) patients. The allele frequency of the G variant of the Q223R polymorphism of the LEPR gene was 79.5% in patients with normal weight and 82.7% in patients with obesity and corresponded to the Hardy-Weinberg equilibrium (p = 0.415 and p = 0.205, respectively). Analysis of genotypes showed a high frequency of genotypes GG - 64.2% and GA - 30.5% in the group with normal BMI and GG - 69.7% and GA - 25.9% in the group with high BMI.

**Table 1.**
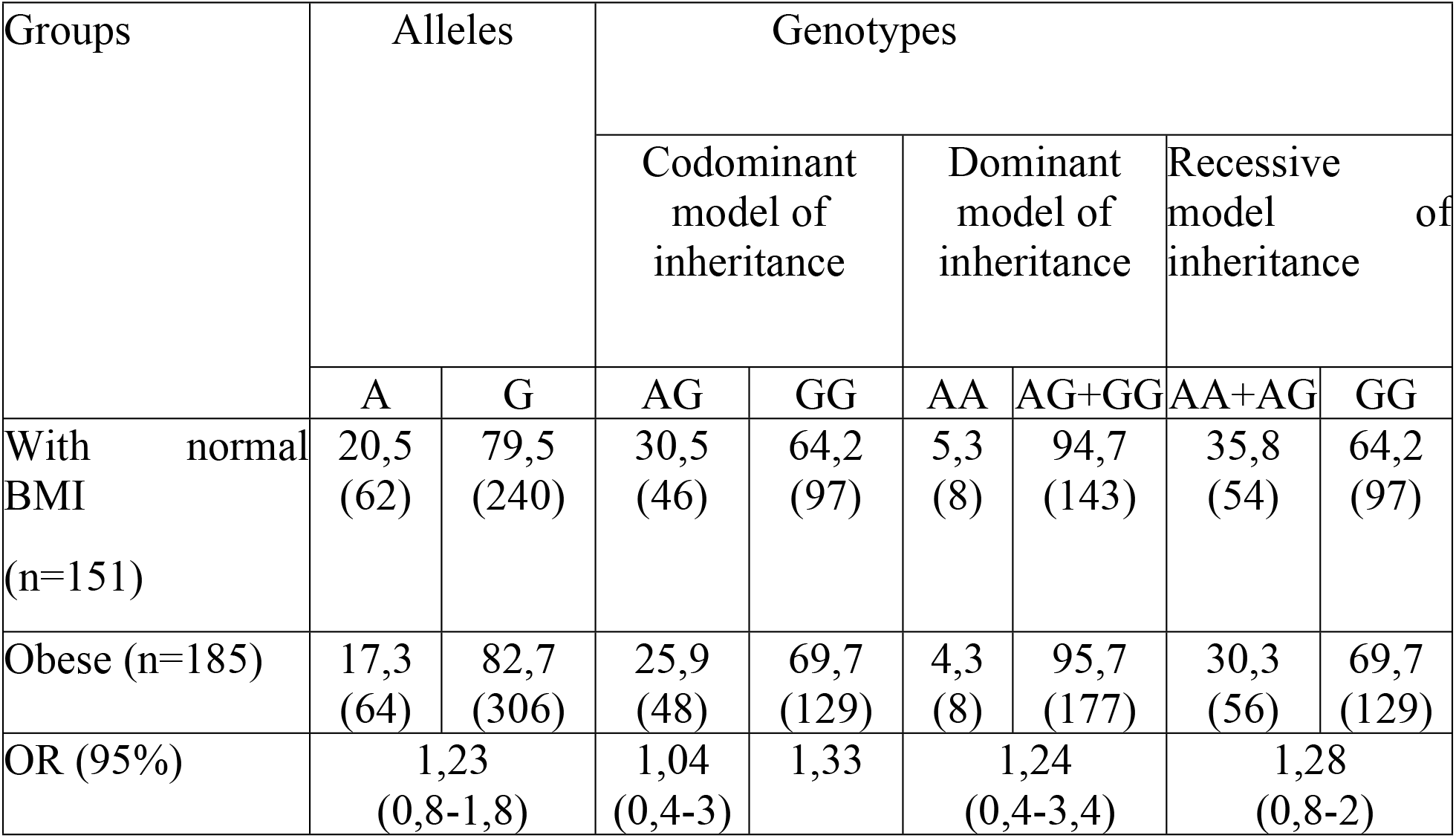
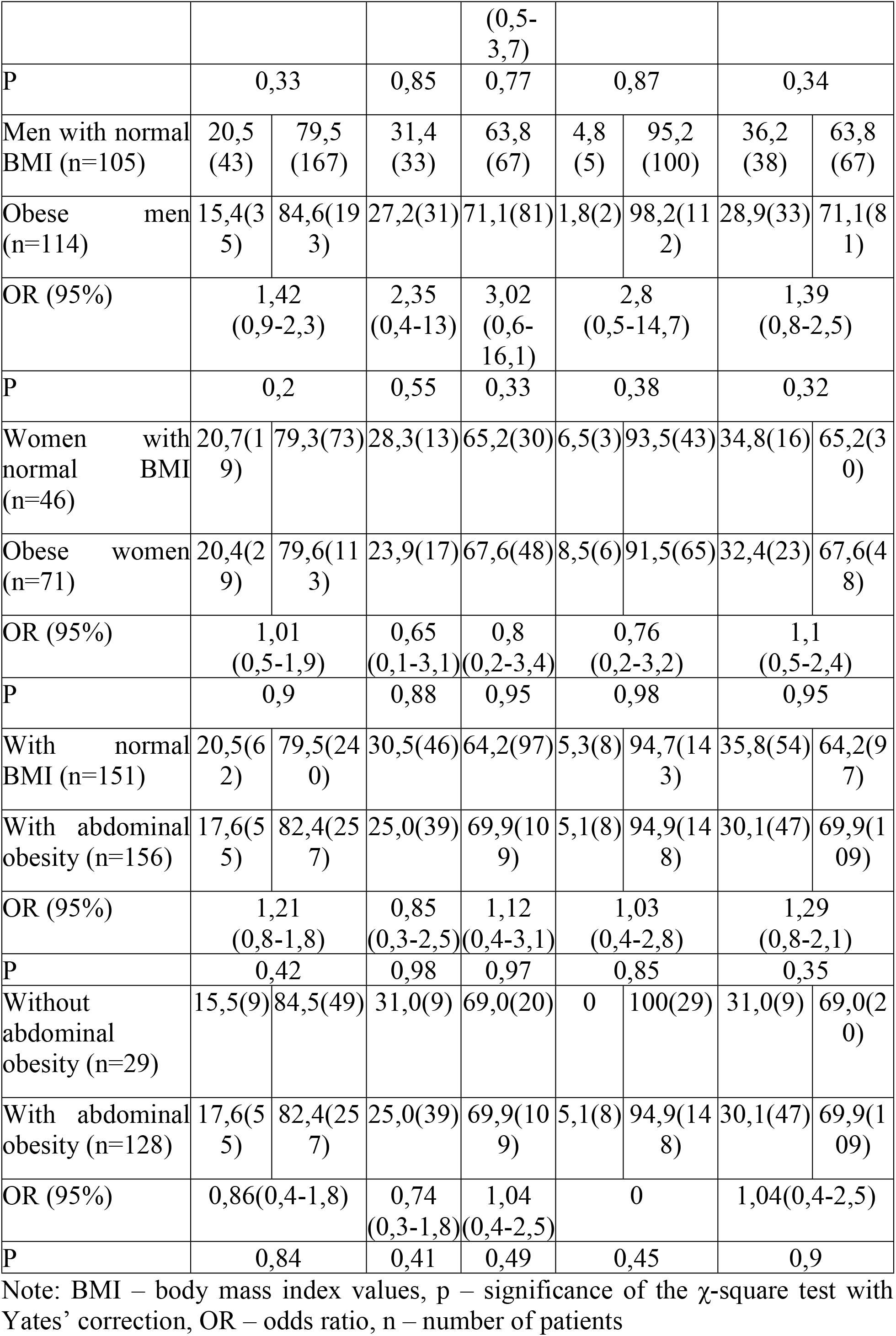
Frequency distribution of alleles and genotypes of the *rs1137101* polymorphism of the *LEPR* gene with odds ratio (OR)

The frequency of alleles and genotypes does not differ in the sample of Yakuts with normal BMI and Yakuts with high BMI. There are also no differences in the frequency of alleles and genotypes based on gender and the presence of abdominal obesity. Analysis of the association between *Q223R LEPR* and obesity in the normal BMI sample and in the obese sample is presented in Table 2.

**Table 2.** Average BMI values depending on the *Q223R* genotype of the *LEPR* gene.

| Genotypes | n | BMI | U | p |
| --- | --- | --- | --- | --- |
| Obese |  |  |  |  |
| AA | 8 | 34 ± 1,22 | 215,5 (*AA/AG*) | >0,05 |
| AG | 48 | 33,4 ± 0,15 | 3063 (*AG/GG*) | >0,05 |
| GG | 129 | 33,1 ± 0,07 | 601 (*AA/GG*) | >0,05 |
| With normal BMI |  |  |  |  |
| AA | 8 | 22,1 ± 0,69 | 145 (*AA/AG*) | >0,05 |
| AG | 46 | 22,7 ± 0,18 | 2128 (*AG/GG*) | >0,05 |
| GG | 97 | 22,6 ± 0,12 | 332,5 (*AA/GG*) | >0,05 |
| With abdominal obesity |  |  |  |  |
| AA | 8 | 34 ± 1,22 | 161,5 (*AA/AG*) | >0,05 |
| AG | 39 | 33,9 ± 0,17 | 2247 (*AG/GG*) | >0,05 |
| GG | 109 | 33,5 ± 0,08 | 475 (*AA/GG*) | >0,05 |

Note: BMI – body mass index values, n – number of patients, U – Mann Whitney test, p – significance of the Mann Whitney test

When analyzing the average anthropometric values depending on the genotype, it is noted that the BMI in carriers of the heterozygous AG genotype was greater than in carriers of the GG genotype. Moreover, in the obese sample, carriers of the AA genotype had the highest BMI. However, differences in BMI indicators depending on genotype in all samples were not statistically significant.

*LEPR* emerged as the most promising candidate gene, likely undergoing natural selection, represented by three variants (*rs1137100, rs1137101*, and *rs1805096*) that strongly distinguish East Asian populations from all other populations described in the literature Table 3.

**Table 3.** Frequency of alleles and genotypes of the *rs1137101* polymorphism of the *LEPR* gene in the Yakut population and in the populations of the 1000 Genomes project.

| Populations | Alleles (%) |  | Genotypes (%) |  |  | Chi-square | p |
| --- | --- | --- | --- | --- | --- | --- | --- |
|  | A | G | AA | AG | GG |  |  |
| Yakutia (this study) | 20,5 | 79,5 | 5,3 | 30,5 | 64,2 | - | - |
| Average frequency in the world | 41,6 | 58,4 | 19,6 | 43,8 | 36,5 | 51,52 | 0,000 |
| South and Middle America | 56,3 | 43,7 | 29,7 | 53,3 | 17,0 | 107,39 | 0,000 |
| Europe | 53,1 | 46,9 | 30,2 | 45,7 | 24,1 | 97,92 | 0,000 |
| South Asia | 49,7 | 50,3 | 25,6 | 48,3 | 26,2 | 78,97 | 0,000 |
| Africa | 40,8 | 59,2 | 15,7 | 50,1 | 34,2 | 42,35 | 0,000 |
| East Asia | 13,1 | 86,9 | 1,6 | 23,0 | 75,4 | 9,6 | 0,002 |

Note: Chi-square – chi – square with Yates correction, p – significance of differences in allele frequencies between populations and the allele frequency of this study.

The high frequency of the G allele in the Yakut population is close to that observed in populations of East Asia (86.9%). In other populations of Europe, Africa, America and South Asia, the G allele occurs with a frequency of 43.7% to 59.2% [13]. Interestingly, the frequency of the G allele in populations of South and Middle America (43.7%) is similar to populations of Europeans (46.9%) and South Asians (50.3%). These data do not fit into modern views that the Indian population and the populations of East Asia (86.9%) have the same roots.

## Discussion

Previously, several studies of *Q223R* and *K109R* polymorphisms in the Yakut population were carried out, so Ammosova E.P. et al. investigated the relationship between the *rs1137100* polymorphism of the *LEPR* gene and the lipid spectrum, metabolic syndrome and its components in the Yakuts. They found that the frequency of the G allele was 66.6%. No association with metabolic syndrome and its components has been identified [5]. Asekritova A.S. et al. conducted a study of *rs1137101* polymorphism in Yakuts, as a result, the frequency of the G allele in patients with metabolic syndrome was 87% and in patients without metabolic syndrome 91% [6]. Ievleva K.D. came to significant results, she found that the risk markers for the implementation of disorders of carbohydrate and energy metabolism against the background of overweight and obesity in Caucasian adolescents (Russians) is the carriage of alleles of the polymorphic loci of the *LEPR* gene *rs1137101* and *rs1137100*, in Mongoloid adolescents (Buryats) - carriage of the *FTO rs9939609* and *FTO rs8050136* alleles [7]. A study in the Malaysian population did not reveal an association between the *rs1137101* and *rs1137100* polymorphisms and obesity [8]. Okada T. et al found an association between the A allele and the incidence of childhood obesity and overweight in the Japanese population [9]. The GENYAL study in Spain assessed 11 SNPs associated with high body mass index in children and found a significant association between *Q223R LEPR* and high weight gain [10].

In their work, the researchers observed a north-south gradient in *Q223R* allele frequency in Europeans, with a higher frequency of derived alleles in the north and a lower frequency in the south. The same phenomenon for SNPs of other genes was reported in the Framingham Heart Study [11] and in a pan-European analysis [12]. In a study of residents of Sri Lanka, Illangasekera Y. A. et al. found a connection between the G allele and BMI and waist circumference; they also found that living in an urban area neutralized the protective effect of the non-risk genotype (AA) in the development of obesity [13].

Our studies in the Yakut population show a high prevalence of variants of the *PNPLA3* and *FABP2* genes associated with increased BMI and non-alcoholic fatty liver disease [15]. In a study by Simcox J. et al, they found that the G allele of the *PNPLA3* gene is associated with adaptation to cold [16]. The authors suggest that the sharply continental climate and specific diet were probably the reason for the high prevalence of variants of genes involved in the metabolism of lipids and carbohydrates, and the accumulation of risk alleles is a consequence of adaptation to living conditions in Yakutia.

*LEPR* is involved in fat storage and heat dissipation by mitochondria, as well as body weight regulation. Polymorphisms in the *LEPR* gene that exhibit some signatures of selection in East Asian populations have been reported to be involved in specific metabolic patterns and/or disorders. For example, *rs1137100* is responsible for a nonsynonymous substitution (*K109R*) that is found to be associated with an increased respiratory quotient (i.e., increased basal metabolic rate), consistent with its important effect on nonshivering thermogenesis.

There is some evidence of adaptation to cold in populations ancestral to anatomically modern humans. Sazini et al. analyzed genes associated with the function of brown adipose tissue, modifications of which contribute to increased heat dissipation by mitochondria, in the genomes of modern populations of East Asia and Europe, as well as in the genomes of fossil hominids (Neanderthals and Denisovans), and found evidence of positive selection for three SNPs in the *LEPR* gene in East Asians [17]. The G variant of the *LEPR* gene (*rs1137101*), showing signs of positive selection, was found in the Neanderthal and Denisovan genomes, suggesting the evolution of independent mechanisms of adaptation to thermal efficiency in these fossil hominin populations. In fact, the variation surrounding *LEPR rs1137100* appears to have actually been shaped by positive selection in East Asian populations, whereas only the potentially cold-adapted *LEPR rs1137101* was observed in archaic species. This suggests that convergent evolution of modern and archaic increased thermogenesis mediated through brown adipose tissue or introgression of related archaic cold-adapted alleles into modern genomes is unlikely [18].

Long-term consumption of fructose has also been shown to lead to leptin resistance. Recently, leptin was found to be associated with autophagy. Autophagy has been demonstrated to be involved in several interesting processes, such as fat storage in adipocytes and liver [19].

The mutant allele has also been reported to reduce leptin inhibition of insulin, leading to insulin dysregulation and increased insulin release, thereby accelerating glucose uptake and basal metabolic rate. Accordingly, this variant could potentially be detrimental in hot climates, in which it did maintain a low frequency, becoming increasingly beneficial in colder climates due to the associated increase in basal metabolic rate and thus heat dissipation, which is the basis of non-shivering thermogenesis. It appears that changes in the *LEPR* gene (*rs1137101* and *rs1137100*) were an advantage for Asian populations 6000–8000 years ago, a time that corresponds to the introduction of agriculture in Asia. It has been suggested that the *LEPR* gene may be considered a “thrifty” gene, leading to the accumulation of adipose tissue in times of plenty, providing a reserve in times of famine. As an alternative explanation for the positive selection of *LEPR* in Asian populations, Hancock et al found associations of several *LEPR* variants with climate variables, suggesting a role for climate adaptation in the biological processes underlying cold adaptation and overweight. They suggest that variants such as *Q223R* and *K109R* may be harmful in hot equatorial climates and beneficial in colder climates [20].

## Conclusions

The authors’ study demonstrates that in the Yakut population, SNP *Q223R* (*LEPR*) is possible and has some effect on anthropometric indicators, but differs from studies conducted on samples of European ethnicity. It can be assumed that the accumulation of the G allele of the *Q223R* (*LEPR*) polymorphism in the Yakut population, as in East Asian populations, is probably a consequence of metabolic adaptation to living conditions in a non-tropical climate.

## Data Availability

All data obtained in this study are available upon reasonable request from the authors.
All data obtained in this work are contained in the manuscript.
All available data is available, except for bioresource collection data containing confidential information

## Acknowledgment

The study was carried out within the framework of the Scientific - Research Experimental - Design Work “Study of the genetic structure and burden of hereditary pathology of populations of the Republic of Sakha (Yakutia)” using the Unique scientific equipment (USE) “The Genome of Yakutia” (No. USU_507512).

## References

1. Tiwari A., Balasundaram P. Public Health Considerations Regarding Obesity. StatPearls. 2023; Available from: https://pubmed.ncbi.nlm.nih.gov/34283488/.

2. Bender N., Allemann N., Marek D., Vollenweider P., Waeber G., Mooser V., Egger M., Bochud M. Association between Variants of the Leptin Receptor Gene (LEPR) and Overweight: A Systematic Review and an Analysis of the CoLaus Study. PLoS One. 2011. 6. e26157. 10.1371/journal.pone.0026157.

3. Hastuti P., Zukhrufia I., Padwaswari M. H., Nuraini A., Sadewa A. H. Polymorphism in leptin receptor gene was associated with obesity in Yogyakarta, Indonesia. Egyptian Journal of Medical Human Genetics. 2016. Vol. 17. Is. 3. P. 271–276. doi: 10.1016/j.ejmhg.2015.12.011

4. Yang Y, Niu T. A meta-analysis of associations of LEPR Q223R and K109R polymorphisms with Type 2 diabetes risk. PLoS One. 2018. Vol. 13 Is. 1. P. e0189366. doi: 10.1371/journal.pone.0189366.

5. Ammosova E.P., Klimova T.M., Zakharova R.N., Fedorov A.I., Baltakhinova M.E. Polymorphic marker rs137100 of the LEPR gene and metabolic disorders in the indigenous population of Yakutia. Yakut Medical Journal. 2021. 2(74). P.5–9 doi 10.25789/YMJ.2021.74.01.

6. Asekritova A. S., Borisova E. P., Kylbanova E. S., Maksimova N. R. Genetic aspects of metabolic syndrome in the Yakut ethnic group. Yakut Medical Journal. 2014. 2 (46). pp. 32–35.

7. Ievleva K.D. Patterns of changes in energy metabolism and the mechanism of its genetic determination in adolescents of two ethnic groups with excess body weight. dis. Ph.D. honey. Sci. Irkutsk, 2022. 138 p.

8. Fan, S.H., Say Y.H. Leptin and leptin receptor gene polymorphisms and their association with plasma leptin levels and obesity in a multi-ethnic Malaysian suburban population. J Physiol Anthropol. 2014. Vol. 33. P.15 doi: 10.1186/1880-6805-33-15.

9. Okada T, Ohzeki T, Nakagawa Y, Sugihara S, Arisaka O, Study Group of Pediatric Obesity and Its related Metabolism. Impact of leptin and leptin-receptor gene polymorphisms on serum lipids in Japanese obese children. Acta Paediatr. 2010. Vol. 99 Is. 8. P. 1213–7. doi: 10.1111/j.1651-2227.2010.01778.x.

10. Marcos-Pasero H, Aguilar-Aguilar E, Colmenarejo G, Ramírez de Molina A, Reglero G, Loria-Kohen V. The Q223R Polymorphism of the Leptin Receptor Gene as a Predictor of Weight Gain in Childhood Obesity and the Identification of Possible Factors Involved. Genes (Basel). 2020. Vol. 11. Is. 5. P. 560. doi: 10.3390/genes11050560.

11. Sebro R, Hoffman T.J., Lange C., Rogus J.J., Risch N.J. Testing for non-random mating: evidence for ancestry-related assortative mating in the Framingham heart study. Genet Epidemiol. 2010. Vol. 34. Is. 7. P. 674–679. doi: 10.1002/gepi.20528.

12. Novembre J, Johnson T, Bryc K, Kutalik Z, Boyko A.R., Auton A, Indap A, King K.S., Bergmann S, Nelson M.R., Stephens M, Bustamante C.D. Genes mirror geography within Europe. Nature. 2008.Vol. 456 Is. 7218. P. 98–101. doi: 10.1038/nature07331.

13. Illangasekera, Y.A., Kumarasiri, P.V.R., Fernando, D.J. et al. Association of the leptin receptor Q223R (rs1137101) polymorphism with obesity measures in Sri Lankans. BMC Res Notes. 2020. Vol. 34 Is. 13. doi: 10.1186/s13104-020-4898-4.

14. GSR and the 1000 Genomes Project. Available from: https://www.internationalgenome.org/

15. Pavlova N.I., Krylov A.V., Alekseev V.A., Bochurov A.A. Association of the Ala54Thr polymorphism of the FABP2 gene with obesity in the Yakut population. Modern problems of science and education. 2023. No. 2.; doi:10.17513/spno.32482.

16. Simcox J., Geoghegan G., Maschek J.A., Bensard C.L., Pasquali M., Miao R., Lee S., Jiang L., Huck I., Kershaw E.E., Donato A.J., Apte U., Longo N., Rutter J., Schreiber R., Zechner R., Cox J., Villanueva C.J. Global Analysis of Plasma Lipids Identifies Liver-Derived Acylcarnitines as a Fuel Source for Brown Fat Thermogenesis. Cell Metab. 2017. Vol. 3 Is. 26. P. 509–522 doi: 10.7554/eLife.52558.

17. Sazzini M, Schiavo G, De Fanti S, Martelli PL, Casadio R, Luiselli D. Searching for signatures of cold adaptations in modern and archaic humans: hints from the brown adipose tissue genes. Heredity (Edinb). 2014. Vol. 113. Is. 3. P. 259–267. doi: 10.1038/hdy.2014.24.

18. Silvert M, Quintana-Murci L, Rotival M. Impact and Evolutionary Determinants of Neanderthal Introgression on Transcriptional and Post-Transcriptional Regulation. Am J Hum Genet. 2019. Vol. 104. Is. 6. P. 1241–1250. doi: 10.1016/j.ajhg.2019.04.016.

19. Aijälä M, Malo E, Ukkola O, Bloigu R, Lehenkari P, Autio-Harmainen H, Santaniemi M, Kesäniemi YA. Long-term fructose feeding changes the expression of leptin receptors and autophagy genes in the adipose tissue and liver of male rats: a possible link to elevated triglycerides. Genes Nutr. 2013. Vol. 8. Is. 6. P. 623–635. doi: 10.1007/s12263-013-0357-3.

20. Bender N, Allemann N, Marek D, Vollenweider P, Waeber G, Mooser V, Egger M, Bochud M. Association between variants of the leptin receptor gene (LEPR) and overweight: a systematic review and an analysis of the CoLaus study. PLoS One. 2011. Vol. 6. Is. 10. P. e26157. doi: 10.1371/journal.pone.0026157.

